# SARS-CoV-2 viremia precedes an IL6 response in severe COVID-19 patients: results of a longitudinal prospective cohort

**DOI:** 10.1101/2022.01.24.22269734

**Authors:** Emilia Roy-Vallejo, Laura Cardeñoso, Ana Triguero-Martínez, Marta Chicot Llano, Nelly Zurita, Elena Ávalos, Ana Barrios, Julia Hernando, Javier Ortiz, Sebastián C. Rodríguez-García, Marianela Ciudad Sañudo, Celeste Marcos, Elena García Castillo, Leticia Fontán García-Rodrigo, Begoña González, Rosa Méndez, Isabel Iturrate, Ancor Sanz-García, Almudena Villa, Ana Sánchez Azofra, Begoña Quicios, David Arribas, Jesús Álvarez Rodríguez, Pablo Patiño, Marina Trigueros, Miren Uriarte, Alexandra Martín-Ramírez, Cristina Arévalo Román, José María Galván-Román, Rosario García-Vicuña, Julio Ancochea, Cecilia Muñoz Calleja, Elena Fernández-Ruiz, Rafael de la Cámara, Carmen Suárez Fernández, Isidoro González Álvaro, Diego A. Rodríguez Serrano, on behalf of the PREDINMUN- COVID Group

## Abstract

**Background:** Interleukin 6 (IL6) levels and SARS-CoV-2 viremia have been correlated with COVID-19 severity. The association over time between them has not been assessed in a prospective cohort. Our aim was to evaluate the relationship between SARS-CoV-2 viremia and time evolution of IL6 levels in a COVID-19 prospective cohort.

**Methods:** Secondary analysis from a prospective cohort including COVID-19 hospitalized patients from Hospital Universitario La Princesa between November 2020 and January 2021. Serial plasma samples were collected from admission until discharge. Viral load was quantified by Real-Time Polymerase Chain Reaction and IL6 levels with an enzyme immunoassay. To represent the evolution over time of both variables we used the graphic command *twoway* of Stata.

**Results:** A total of 57 patients were recruited, with median age of 63 years (IQR [53-81]), 61.4% male and 68.4% caucasian. The peak of viremia appeared shortly after symptom onset in patients with persistent viremia (more than 1 sample with >1.3 log10 copies/ml) and also in those with at least one IL6>30 pg/ml, followed by a progressive increase in IL6 around 10 days later. Persistent viremia in the first week of hospitalization was associated with higher levels of IL6. Both IL6 and SARS-CoV-2 viral load were higher in males, with a quicker increase with age.

**Conclusions:** In those patients with worse outcomes, an early peak of SARS-CoV-2 viral load precedes an increase in IL6 levels. Monitoring SARS-CoV-2 viral load during the first week after symptom onset may be helpful to predict disease severity in COVID-19 patients.

## 1 Introduction

One of the most feared complications of the disease caused by the coronavirus SARS-CoV-2 (COVID-19), is the development of an Acute Respiratory Distress Syndrome (ARDS), which can affect 15.6-31% of patients [1]. Siddiqui and Mehra [2] proposed that ARDS is part of the final stage of the disease, in which clinical features are mainly the consequence of the host hyperinflammatory response and a cytokine storm; whereas the stage I (early infection) is mainly caused by viral replication and the early immune response. However, to the best of our knowledge, this proposal has not been validated.

Since the outbreak of the COVID-19 pandemic, many efforts have been made to find early risk factors and biomarkers able to predict the evolution towards the cytokine storm. In this sense, older age, obesity and comorbidities such as hypertension, diabetes and coronary heart disease have been associated with higher risk of death [3,4]. On the other hand, increased levels of C reactive protein (CRP), lactate dehydrogenase (LDH) and D-dimer, among others, have been shown to be related to the development of ARDS and mortality [3,5,6].

In this context, Interleukin 6 (IL6) has been described as one of the most useful biomarkers [7]. In a previous work, we showed that IL6 could be a severity biomarker but also a guide to select COVID-19 patients who could benefit from treatment with tocilizumab, an inhibitor of the IL6 receptor [8]. Another important biomarker is the presence of SARS-CoV-2 RNA in peripheral blood (viremia), which has been associated with disease severity and a hyperinflammatory state [9,10]. Saji et al.[11] showed that the combination of SpO2/FiO2, IL6 and the presence of SARS-CoV-2 viremia at admission had the highest accuracy to predict fatal outcomes. Bermejo et al. [12] and Myhre et al.[13] found that the presence of SARS-CoV-2 viremia at admission correlated with increased levels of IL6, CRP and ferritin. In addition, a proteomic analysis showed that the expression of viral response and interferon/monocytic pathway proteins such as IL6 and one of its regulators, the Nicotinamide phosporibosyl transferase (NAMPT), were upregulated in patients with quantifiable SARS-CoV-2 viremia at admission, compared to those with undetectable viremia [14].

In a previous study of our group, we found that viremia was associated with Intensive Care Unit (ICU) admission and in-hospital death, and it was a better biomarker than IL6 [10]. In this regard, since SARS-CoV-2 infection is involved in triggering IL6 expression, viremia as an indicator of the systemic viral shedding, could be related with the IL6 response and be useful as an early biomarker (phase of viral response). Nevertheless, the factors determining an IL6 increase in COVID-19 patients have not been well established yet and the association over time between SARS-CoV-2 viremia and IL6, has not been assessed in a prospective cohort with serial samples.

Considering our previous results, the aim of this study was to evaluate the relationship between the presence of SARS-CoV-2 viremia and the time evolution and IL6 levels in a prospective cohort of COVID-19 hospitalized patients.

## 2 Materials and Methods

### 2.1 Study design, population and data collection

This work is a secondary analysis of samples from a prospective observational cohort assembled to validate the predictive value of SARS-CoV-2 viremia (ongoing manuscript). The study included patients hospitalized for COVID-19 in Hospital Universitario La Princesa (HUP) between November 1st 2020 and January 15th 2021.

The inclusion criteria were: a) positive Real-Time Reverse Transcription Polymerase Chain Reaction (rRT-PCR) for SARS-CoV-2 in nasopharyngeal and throat swabs at most 48 hours prior to hospitalization; b) acceptance to participate in the study and oral or written consent; c) age older than 18 years. The exclusion criteria were: a) patients without an initial viremia determination in the first 24-36 hours after admission; b) patients unlikely to be followed-up because they were candidates to be transferred to other health facilities.

Clinical, laboratory and therapeutic data were collected from electronic clinical records and included in an anonymized database. Baseline clinical and laboratory data are to those obtained at admission day.

The need for hospitalization was decided by the physicians at the emergency room based on clinical criteria, without the intervention of the research team. Patient’s treatment and management was decided by each attending physician based on the hospital protocols and their clinical judgment. Attending physicians were blind to the viremia results.

### 2.2 Sample size

The sample size was estimated in 49 patients to validate the primary objective of the study “SARS-CoV-2 viremia as a biomarker of disease severity” (ongoing manuscript) based on the results of our previous retrospective studies [10,15].

### 2.3 Sample collection

Serial plasma and serum samples were collected from admission until discharge. In the first week, samples were collected every 48 hours, with the first sample in the first 24-36h. Thereafter, samples were collected twice a week. All samples were frozen at -80 ºC and stored in the Microbiology Department facilities.

### 2.4 SARS-CoV-2 RNA extraction and detection

Firstly, a nucleic acid extraction of samples was performed by the automatic eMAG® Nucleic Acid Extraction System (Biomerieux, France). Detection of viremia was performed with rRT-PCR using TaqPath™ COVID-19 CE81 IVD RT-PCR Kit (Thermo Fisher Scientific, USA [TFS]), according to the manufacturer’s instructions, by a QuantStudio™ 5 Real Time PCR System (Applied Biosystems, USA). Amplification curves were analyzed with QuantStudio™ Design and Analysis software version 2.4.3 (Applied Biosystems, USA) and interpreted by a clinical microbiologist. To increase sensitivity, two wells were used for each sample. Two positive controls (one of 20,000 copies and another of 200 copies) and two negative controls were added in each run in duplicate.

### 2.5 Quantification of viral load

A standard curve was established using a positive control with a known concentration (TaqPath Positive Control from TaqPath™COVID-19 Control Kit) of 10,000 copies/μl of the SARS-CoV-2 genomic regions targeted by the TFS assay. Ten-fold serial dilutions of the positive control were made up to 1 copy. Nine wells of each of the concentrations and of the negative control were added to the run. The rRT-PCR was performed by QuantStudio™ 5 Real Time PCR System and a standard curve was obtained plotting DNA concentration against cycle threshold (Ct) values. The amplification curves were analyzed with QuantStudio™ Design and Analysis software version 2.4.3 (Applied Biosystems, USA). The results of the nine wells with 1 copy were omitted because they were widely dispersed. Viral load was calculated from Ct values using the standard curve as reference and was expressed as copies/ml and the logarithm with base 10 (log10). Due to the variability and lack of accuracy obtained with the lower levels of SARS-CoV-2 viremia, only viremias >1.3 log10 (20 copies/ml) were considered quantifiable.

### 2.6 IL-6 measurement

Serum samples collected in the same extraction as the plasma used for SARS-CoV-2 viremia determinations were used to assess IL6 levels. IL6 levels were retrospectively quantified in triplicate with the Human IL6 Duoset enzyme-immunoassay from R&D Systems Europe Ltd (Abingdon, UK), following the manufacturer’s instructions.

### 2.7 Variables

For analysis using viremia as a quantitative variable, all values were used. However, to define viremia as cathegorical, positive viremia was considered when values were higher than 1.3 log10 (namely 32 copies/ml, which was the threshold for quantifiable viremia) and negative when values were below this threshold. Persistent viremia was defined as more than one positive viremia in the first week of hospitalization.

Two different variables were used to evaluate IL6 levels: a) a quantitative variable defined as IL6 concentration, expressed in pg/ml, b) a dichotomic variable, which considered levels of IL6 as high when at least one IL6 determination was higher than 30 pg/ml or low if all determinations were below 30 pg/ml. This threshold was based on our previous study, where we showed that IL6>30 pg/ml was associated with poor respiratory outcomes [8]. The average levels of IL6 and viral load were defined as the arithmetic mean of all their determinations in each patient.

### 2.8 Statistical analysis

We used Stata 14.0 for Windows (Stata Corp LP, College Station, TX, USA) for all the analysis described below. Quantitative variables were represented as median and Interquartile Range (IQR), and the Mann Whitney or Kruskall Wallis tests were used to assess significant differences, since all quantitative variables followed a non-normal distribution. Qualitative variables were described as counts and proportions and Chi square or Fisher’
ss exact test was used for comparisons.

In order to comparatively show levels of IL6 and viral load through the two first weeks of follow-up, we used as time variable the number of days from the beginning of symptoms to collection of each sample. To represent the mean evolution over time of both variables we used the graphic command *twoway* from Stata with the option fractional polynomial prediction with 95% confidence interval (CI). Since it is well known that blockade of IL6 receptor with tocilizumab can result in an increase of IL6 serum levels [16], we decided to carry forward the last observation before tocilizumab treatment (last observation carried forward [LOCF] strategy) to replace IL6 values in the remaining visits of the first 2 weeks for those patients treated with tocilizumab in order to avoid the bias of excluding this important population (see comparative baseline characteristics in Suplementary Table 1). Furthermore, we also applied LOCT strategy for those patients who died or were discharged before the 5^th^ visit (14^th^ day after admission), in order to obtain a more homogeneous number of determinations all along the follow-up.

To determine which variables were associated with high levels of IL6, we performed a multivariable logistic regression analysis that was first modelled by adding all the variables with a p value lower than 0.15 in the bivariable analysis. The final model was reached through backward stepwise removal of variables with p value higher than 0.15.

### 2.9 Ethics

This study was approved by the Research Ethics Committee of Hospital Universitario La Princesa, Madrid, (register number 4267; 22-10-2020) and it was carried out following the ethical principles established in the Declaration of Helsinki. As proposed by AEMPS (Agencia Española de Medicamentos y Productos Sanitarios, The Spanish Agency for Medicines and Medical Devices), only oral consent was required due to the COVID-19 emergency [17]. However, a written information sheet was also offered to all patients. After being informed about the study, all included patients (or their representatives) gave informed consent, which was registered in the electronic clinical chart.

This article was written following the STROBE initiative (Strengthening the Reporting of Observational studies in Epidemiology).

## 3 Results

### 3.1 Study population and sample characteristics

A total of 57 patients were recruited, with median age of 63 years (IQR 53-81), 61.4% were male, 68.4% caucasian, and 75.4% had previous comorbidities. The median time from symptom onset to first sample was 8 days (IQR 4-10). Baseline clinical characteristics according to IL6 levels are shown in table 1. During patients’ hospitalization, 301 serum samples were collected, with a median number of 3 samples per patient (IQR 2-5).

**Table 1.**
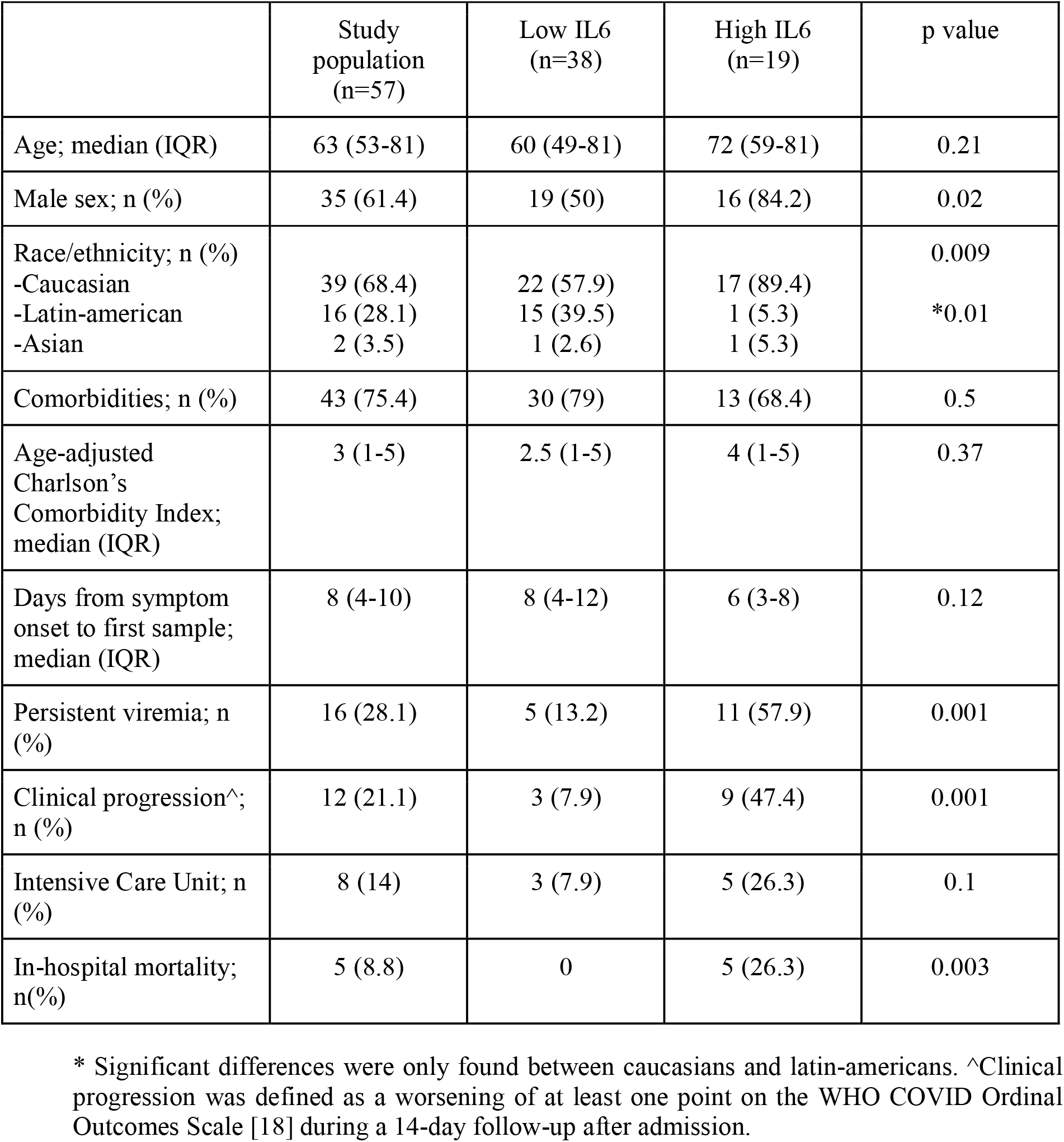
Baseline clinical characteristics of the study population according to IL6 levels.

Nine patients were treated with tocilizumab, who on average, showed data suggesting a more severe disease, although differences did not reach statistical significance (Supplementary Table 1).

Most patients who progressed to a severe disease started this evolution 7 to 14 days after the symptoms onset. In addition, patients with a more benign course were discharged at the end of the first week after admission. For clinical consistency, we decided to analyze only the samples corresponding to the first two weeks of hospitalization, a maximum of 5 samples per patient. Thus, IL6 levels were measured in 228 samples, with a median of 3.6 pg/ml (IQR 0-21 pg/ml). Baseline clinical characteristics of patients depending on IL6 status are shown in table 1. SARS-CoV-2 viremia was determined in 234 samples, with the highest percentage of positive viremia (36.8%) at admission (visit 1).

### 3.2 Time course of IL6 and SARS-CoV-2 viremia

The average serum levels of IL6 and SARS-CoV-2 viral load were moderately but significantly correlated (r=0.41, p=0.0014; Supplementary Fig1).

Figure 1A shows the evolution over time of IL6 and viremia analyzed using data from the whole population (including IL6 after tocilizumab treatment). The peak of viremia appeared early, at the first days after symptom onset (day 3 to 5), and quickly decreased. On the other hand, the highest levels of IL6 were found at day 20. The wide 95% CI suggested a high heterogeineity, specially at both extremes of the time course. Figure 1B shows the results when the LOCF strategy (see Statistical section for further information) was used to minimize the increase of IL6 induced by tocilizumab (see supplementary Figure 2 for raw data in cases treated or not with tocilizumab). With LOCF strategy, the peak of IL6 was smaller, but the time course of IL6 production was quite similar to that obtained from raw data. Hereinafter, the relationship between IL6 and SARS-CoV2 viral load shown corresponds to results obtained with the LOCF strategy.

**Figure 1.**
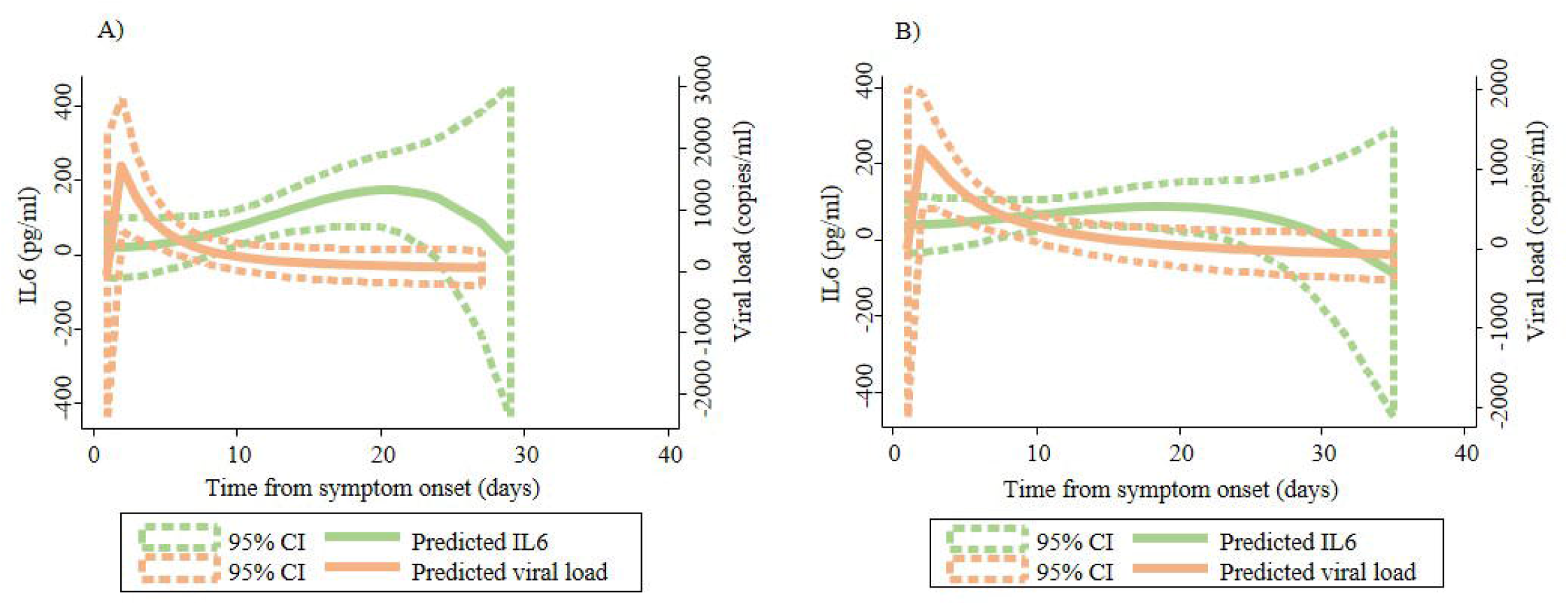
The peak of viral load precedes the IL6 increase. Graphic representation of time-course of IL6 levels and SARS-CoV-2 viral load from symptom onset. Panel (A) representation of raw data. Panel (B) Representation of data after applying the LOCF strategy. The fractional polynomial prediction was performed using the *twoway* command of Stata.

### 3.3 Relationship between IL6 and SARS-CoV-2 viremia

A total of 19 patients had high IL6 (Table 1), of them 11 (57.9%) had persistent viremia compared to 5 patients (13.2%) in the low IL6 group (p=0.001), with an odds ratio of 9.1 (95%CI 2.5-32.6) (Figure 2E). In the graphic representation of IL6 and viral load according to high/low IL6 (Figure 2A, B), an early and minor peak of IL6 was found in the low IL6 group, together with a small peak of viremia at day 4. On the other hand, patients with at least one IL6 above 30 pg/ml had an early high viremia around day 3 and a progressive increase of IL6 especially after day 12.

**Figure 2.**
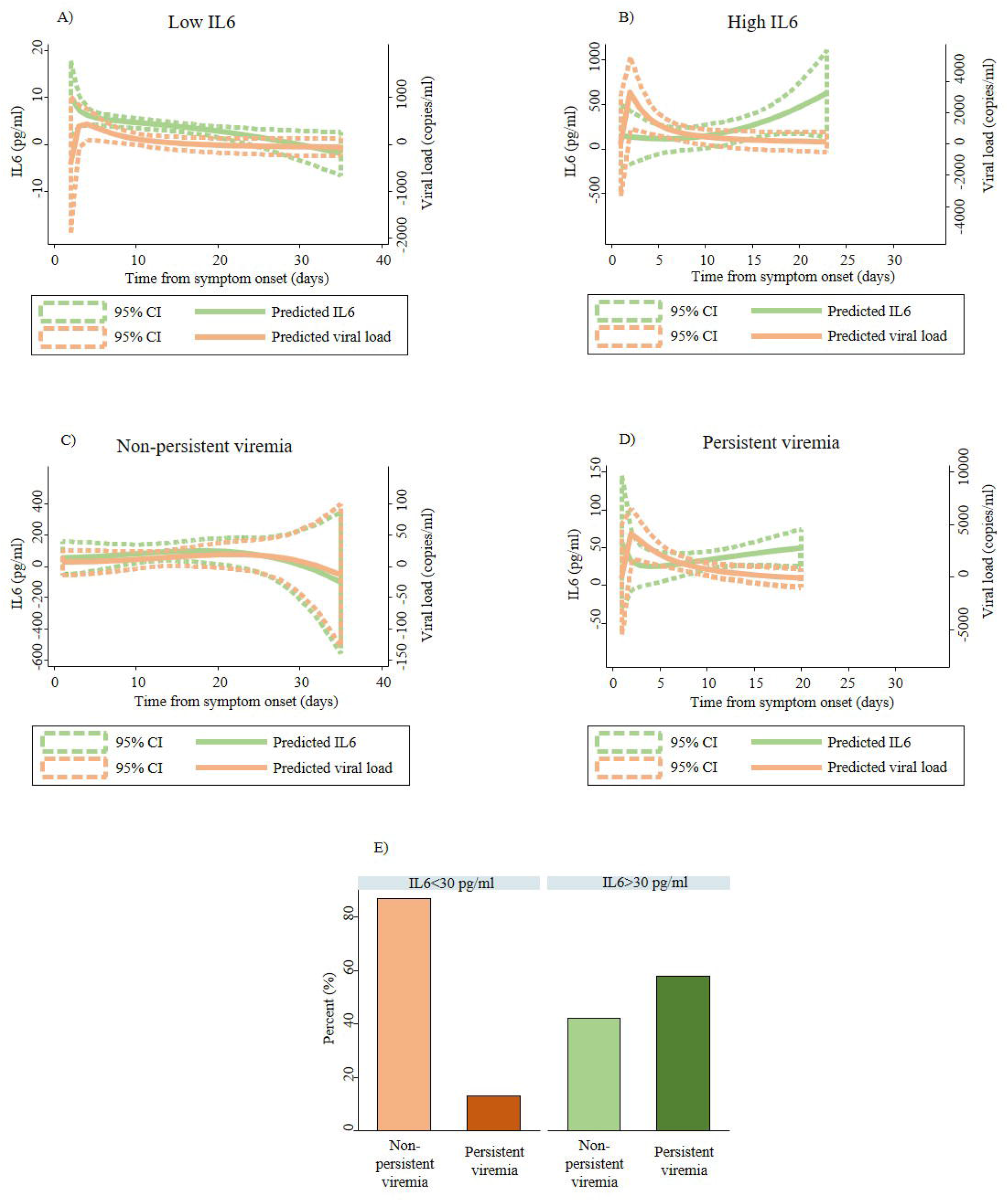
Patients with worse outcomes have an early peak of SARS-CoV-2 viral load before a prominent increase in IL6 levels. Graphic representation of IL6 levels and SARS-CoV-2 viral load from symptom onset in: (A) patients with low IL6; (B) patients with at least one IL6>30 pg/ml (high IL6); (C) non-persistent viremia and (D) persistent viremia. Pannel (E) represents the percentage of patients with persistent viremia according to IL6 levels (low versus high). Panels (A) to (D), the fractional polynomial predictions were performed using the *twoway* command of Stata.

When the prediction of IL6 and viral load was calculated according to persistent viremia status, remarkable differences were obtained (Figure 2C, D). In the persistent viremia group, viral load showed a peak around day 4, whereas IL6 had a two-phase increase: one at the first days from symptom onset and then a subsequent progressive increase after day 5. Regarding non-persistent viremia the increase of IL6 was slow from symptom onset until day 20. The median of the average levels of IL6 were 3.6 pg/ml (IQR 1.0-9.2 pg/ml) in the non-persistent viremia group and 21.4 pg/ml (IQR 12.3-44.9) in patients with persistent viremia (p<0.001).

### 3.4 Prediction of IL6 and SARS-CoV-2 viremia according to demographic factors

The effect of demographic factors on IL6 levels and SARS-CoV-2 viremia was also assessed. The median of the average IL6 concentration was significantly higher in males (11.3 pg/ml [IQR 3.3-27.5] than in females (2.5 pg/ml [IQR 0.7-9.2 pg/ml]; p=0.005). In the group of patients with high IL6, 84.2% were male compared to 50% in the group with low IL6 (p=0.02). No differences were found in the average viral load (11.1 copies/ml [IQR 0-197.3 copies/ml] vs 2.3 copies/ml [IQR 0-6.8 copies/ml]; p=0.08) or in the percentage of patients with persistent viremia (34.3% vs 18.2%; p=0.24) between males and females, respectively. However, predicting curves for IL6 and viral load were substantially different depending on sex (Figure 3A). In males, curves had a fast increase in viral load with a peak around day 2 from symptom onset and a later rise in IL6 levels until day 20, while women showed a small increase in viral load and IL6 between day 2 and 7 approximately.

**Figure 3.**
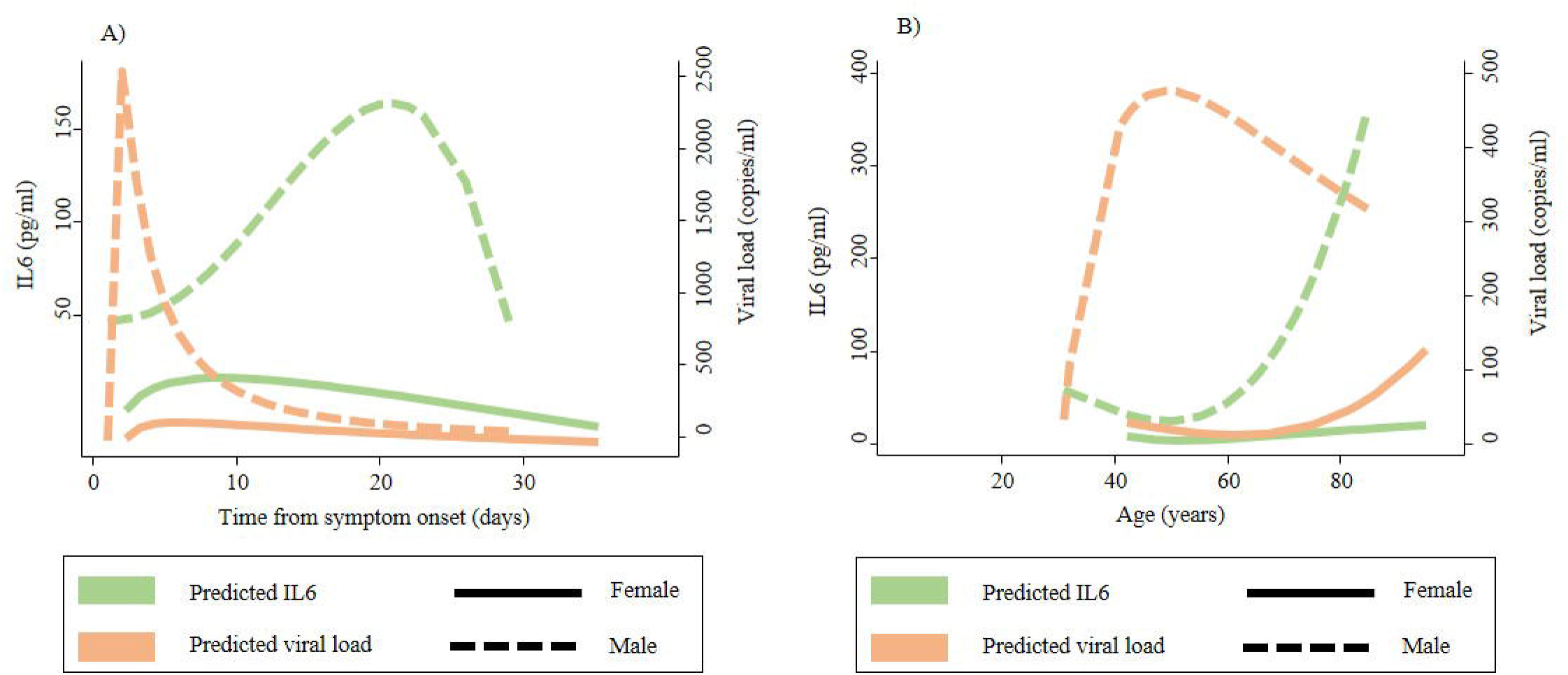
Males had more relevant increases of IL6 and viral load. Panel (A) represents the levels of IL6 and viral load from symptom onset by sex, while panel (B) shows levels of IL6 and viral load by age and sex. The fractional polynomial predictions were performed using the *twoway* command of Stata.

The effect of age on IL6 levels and viral load was also considered. The average levels of IL6 and viremia did not correlate with age (r=0.21, p=0.13; and r=0.19, p=0.16; respectively). Moreover, no differences were found when age was categorized as <75 years and >75 years (p=0.57 and p=0.88, for IL6 and viral load respectively). In the group with high IL6, 31.6% of patients were older than 75 years, the same percentage as in the low IL6 group (p=1). Regarding persistent viremia, the proportion of patients older than 75 years was 29.3 % the group with non-persistent viremia and 37.5% in those with persistent viremia (p=0.55). In the prediction curves according to age and sex, all parameters increased with age except for viral load in males, which peaked between 40 and 50 years (Figure 3B).

Regarding ethnicity, there were only differences between caucasians and latin-americans in the average IL6 levels (11.9 [IQR 2.9-35.2] vs 2.9 [IQR 1.0-4.3]; p= 0.005), and the percentage of patients with high IL6 (89.5% vs 5.3%, p=0.01) (Table 1). No differences were found depending on ethnicity in the average viral load or the percentage of patients with persistent viremia.

Finally, a multivariable analysis showed that the presence of high IL6 was associated with persistent viremia (OR 10.0 [95%CI 2.0-49.5]; p=0.005); conversely, female sex (OR 0.17 [0.03-0.9]; p=0.04) and latin-american origin (OR 0.06 [0.01-0.7]; p=0.02) had a protective effect.

## 4 Discussion

This study assessed the relationship between IL6 and SARS-CoV-2 viremia. The most relevant finding was the different time course of IL6 and viremia: the peak of viremia appeared shortly after symptom onset in patients with persistent viremia and also in those with at least one measure of IL6>30 pg/ml, in which it was followed days later by a progressive increase in IL6. Moreover, the presence of persistent viremia in the first week of hospitalization was associated with higher levels of IL6. Both IL6 and SARS-CoV-2 viral load were higher in males, with a progressive increase with age that occurred earlier in males.

Our findings are consistent with the COVID-19 phases first described by Siddiqui and Mehra [2]. These authors described an early stage characterized by a viral response (SARS-CoV-2 viremia) and a final stage caused by a hyperinflammatory state characterized by increased levels of IL6. To date, the only study that has evaluated the kinetics of SARS-CoV-2 viremia and systemic cytokines, found that the levels of IL6 were associated with critical disease but not with the presence of viremia. However, only 20 patients were included in this study and the authors did not consider the different time course of the increase in viremia and IL6 [19]. In our previous work [10], we described that the presence of relevant SARS-CoV-2 viremia was associated with higher risk of death and ICU admission. Furthermore, viremia was the most useful biomarker for these outcomes, being superior to IL6, lymphopenia and LDH. In the present study, we show that the SARS-CoV-2 viremia appears early in the course of the disease, standing out as a relevant, simple and early biomarker.

Since the beginning of the pandemic, CRP and IL6 levels have not only been used as prognostic biomarkers in COVID-19 but also to guide treatment and predict response to tocilizumab [8,17,20]. However, Ong et al. and Liu et al. [21,22] showed that IL6 in COVID-19 patients peaked after the worsening of respiratory function, suggesting that when proinflammatory biomarkers rise, lung damage might be already established. In our cohort, 68.8% of patients with persistent viremia had at least one IL6> 30 pg/ml in the later hyperinflammatory phase of the disease. Taking into account the high percentage of patients with persistent viremia who develop an hyperinflammatory response, these patients might be considered as candidates for intensive treatment and surveillance, or even for early treatment with IL6 blockade.

Nevertheless, these findings might not be extensive to all patients. A more severe course of COVID-19 and higher levels of IL6 have been previously described in older males [23]. In this sense, genetic and hormonal factors have been proposed to be involved in age and sex related differences in COVID-19 [23,24]. One of the most studied SARS-CoV-2 related proteins is ACE2, the membrane receptor needed for the virus internalization, which is encoded by the gen of the same name located in the X chromosome [24,25]. ACE2 expression increases with age and in male sex in COVID-19 patients [26,27]. ACE2 expression also correlates with SARS-CoV-2 infectivity in cells of the respiratory tract [28] and with higher viral loads in nasopharyngeal swabs [29]. Whether endothelial and vascular ACE2 is related to viral load in peripheral blood has not been assessed yet, but it is plausible that higher levels of systemic ACE2 lead to increased viremia. Another proposed mechanism to explain sex differences is the effect of Toll Like Receptor (TLR) pathways, especially TLR7. This receptor, which recognizes viral single strain RNA and enhances IL6 production, is also located in X chromosome. TLR7 was one of the most important susceptibility genes found in an Italian cohort of COVID-19 patients, where 6.3% of young males with life-threatening disease presented missense variants of this gene[30].

Regarding ethnicity, patients with a latinamerican origin had lower levels of IL6 in our cohort. In this sense, other cytokines and chemokines such as MCP-1, IL-10, IL-15, CXCL10 and CCL2 have been associated with SARS-CoV-2 viremia [12,19]. It is possible that the immune response of latinamerican patients is enhanced by molecular pathways different from IL6, but this hypothesis needs to be further evaluated with studies with bigger sample size of our cohort.

This study has several limitations. First of all, the sample size of our cohort was small, although it was sufficient to find different patterns in the kinetics of IL6 and viral load. Secondly, all patients included were hospitalized and their first sample was obtained at a median of 7 days after symptom onset, therefore, data from the first days of the disease were limited. In addition, information about different variants of SARS-CoV-2 in our cohort could not be obtained because viral sequencing was not available in our facilities. However, at the time our study was performed, the most prevalent variant in Madrid was the original strain [31].

In conclusion, in those patients with worse outcomes, an early peak of SARS-CoV-2 viral load precedes around 5-10 days a prominent increase in IL6 levels. This finding was very clear in males older than 40 years. Therefore, monitoring SARS-CoV-2 viral load during the first week after symptom onset may be helpful to stratify the severity of patients and predict those who are at high risk of developing hyperinflammatory syndrome and ARDS.

## Supporting information

STROBE Checklist

## Data Availability

All data produced in the present study are available upon reasonable request to the authors

## 6 Author Contributions

ER-V, LC, IG-A and DAR-S designed the study and wrote the first draft of the manuscript; MCL, EA, AB, JH, JO, SCR-G, MC, CM, EGC, BG, RM, II, AV, AS-A, DA, JAR, PP, MT, MU, JMG-R, RG-V, JA, RdC and CSF included patients in the study and collected data; BQ, CAR, CM-C and EFR extracted and processed samples; ER-V, AT-M, NZ and LFG-R performed laboratory determinations; ER-V, AS and IG-A analyzed data; all authors reviewed the final draft.

## 8 Acknowledgments

Special thanks to our patients and relatives for agreeing with the use of pseudonymized clinical data and surpluses of clinical samples to perform this study, and Manuel Gomez Gutierrez, PhD for his excellent editing assistance.

## PREDINMUN-COVID group includes

Internal Medicine-Infectious Diseases: Jesús Sanz^1^, Pedro Casado^1^, Ángela Gutiérrez^1^, Azucena Bautista^1^, Pilar Hernández^1^, Nuria Ruiz Giménez^1^, Berta Moyano^1^, Paloma Gil^1^, María Jesús Delgado^1^, Pedro Parra^1^, Beatriz Sánchez^1^, Carmen Sáez^1^, Marta Fernández Rico^1^.

Microbiology: Diego Domingo García^2^, Teresa Alarcón Cavero^2^, María Auxiliadora Semiglia Chong^2^, Ainhoa Gutiérrez Cobos^2^.

Rheumatology: Santos Castañeda^3^, Irene Llorente^3^, Eva G. Tomero^3^, Noelia García Castañeda^3^, Nuria Montes^3^.

Intensive Care Unit: Cristina Dominguez Peña^4^, David Jiménez Jiménez^4^, Pablo Villamayor^4^, Alfonso Canabal^4^.

Pneumology: Tamara Alonso^5^, Carolina Cisneros^5^, Claudia Valenzuela^5^, Francisco Javier García Pérez^5^, Rosa María Girón^5^, Javier Aspa^5^, Celeste Marcos^5^, M. del Perpetuo Socorro Churruca^5^, Enrique Zamora^5^, Adrián Martínez^5^, Mar Barrio Mayo^5^, Rosalina Henares Espi^5^.

Immunology: Francisco Sánchez-Madrid^9^, Enrique Martín Gayo^9^, Ildefonso Sánchez-Cerrillo^9^, Ana Marcos Jimenez^9^, Pedro Martínez-Fleta^9^, Celia López-Sanz^9^, Ligia Gabrie^9^, Luciana del Campo Guerola^9^, Reyes Tejedor^9^, Rosa Carracedo-Rodríguez.

