## Supplementary Material for "SARS-CoV-2 viremia precedes an IL6 response in severe COVID-19 patients: results of a longitudinal prospective cohort"

**1 Supplementary Table**

Supplementary Table 1. Baseline clinical characteristics of the population according to treatment with tocilizumab

|  | Patients not treated with tocilizumab (n=48) | Patients treated with tocilizumab  (n= 9) | p value |
| --- | --- | --- | --- |
| Age; n (IQR) | 63 (50.5-81) | 63 (56-81) | 0.69 |
| Male sex; n (%) | 28 (58.3) | 7 (77.8) | 0.46 |
| Race/ethnicity; n (%)   - Caucasian - Latin-american - Asian | 33 (68.7)  14 (29.2)  1 (2.1) | 6 (66.7)  2 (22.2)  1 (11.1) | 0.37 |
| Comorbidities; n(%) | 36 (75) | 7 (77.8) | 1 |
| Age-adjusted Charlson’s Comorbidity Index | 3.5 (1-5) | 3 (2-4) | 0.37 |
| Days from symptom onset to first sample | 7 (4-10) | 9 (8-12) | 0.28 |
| Persistent viremia | 11 (22.9) | 5 (55.6) | 0.1 |
| Clinical progression | 9 (18.8) | 3 (33.3) | 0.38 |
| Intensive Care Unit | 7 (14.6) | 1 (11.1) | 1 |
| In-hospital mortality | 3 (6.25) | 2 (22.2) | 0.17 |

**2 Supplementary Figures**

0

50

100

150

200

0

1000

2000

3000

4000

5000

Average viral load (copies/ml)

IL6 average levels

95% CI

Fitted values

IL6 (pg/ml)

Supplementary Figure 1. Correlation bertween average levels of IL6 and viral load. The graphic was performed using the command *twoway* of Stata

No treatment with tocilizumab

-600

-400

-200

0

200

400

-100

0

100

200

0

10

20

30

40

95% CI

Predicted IL6

95% CI

Predicted viral load

Time from symptom onset (days)

IL6 (pg/ml)

Viral load (copies/ml)

0

5000

10000

15000

-500

0

500

1000

1500

0

10

20

30

95% CI

Predicted IL6

95% CI

Predicted viral load

Time from symptom onset (days)

IL6 (pg/ml)

Viral load (copies/ml)

Treatment with tocilizumab

A)

B)

Supplementary Figure 2. Graphic representation of IL6 levels and SARS-CoV-2 viral from symptom onset in: A) patients not treated with tocilizumab; B) patients treated with tocilizumab. Panels A and B were performed using the twoway command of Stata.
